# THE IMPACT OF ROUTINE CARDIAC TROPONIN I-BASED CARDIOTOXICITY SCREENING ON CLINICAL OUTCOMES IN PATIENTS ON CANCER IMMUNOTHERAPY

**DOI:** 10.1101/2024.01.22.24301442

**Authors:** Maja Ivanovic, Antonia Chan, Evaline Cheng, Sidra Xu, Carissa Lee, Jonathan You, Miguel Franquiz, Muhammad Fazal, Ryan Batchelder, Sean M. Wu, Sunil A. Reddy, Tamiko Katsumoto, Kavitha Ramchandran, A. D. Colevas, Saad A. Khan, Alice C. Fan, Paul Cheng, Heather Wakelee, Ronald Witteles, Joel W. Neal, Sarah Waliany, Han Zhu

## Abstract

**Purpose:** Immune checkpoint inhibitors (ICI) used as cancer therapy have been associated with a range of cardiac immune-related adverse events (irAEs), including fulminant myocarditis with a high case fatality rate. Early detection through cardiotoxicity screening by biomarker monitoring can lead to prompt intervention and improved patient outcomes. In this study, we investigate the association between cardiotoxicity screening with routine serial troponin I monitoring in asymptomatic patients receiving ICI, cardiovascular adverse event (CV AE) detection, and overall survival (OS).

**Methods:** We instituted a standardized troponin I screening protocol at baseline and with each ICI dose (every 2-4 weeks) in all patients receiving ICI at our center starting Jan 2019. We subsequently collected data in 825 patients receiving ICI at our institution from January 2018 to October 2021. Of these patients, 428 underwent cardiotoxicity screening with serial troponin I monitoring during ICI administration (Jan 2019-Oct 2021) and 397 patients were unmonitored (Jan 2018-Dec 2018). We followed patients for nine months following their first dose of ICI and compared outcomes of CV AEs and OS between monitored and unmonitored patients. Additionally, we investigated rates of CV AEs, all-cause mortality, and oncologic time-to-treatment failure (TTF) between patients with an elevated troponin I value during the monitoring period versus patients without elevated troponin I.

**Results:** We found a lower rate of severe (grades 4-5) CV AEs, resulting in critical illness or death, in patients who underwent troponin monitoring (0.5%) compared to patients who did not undergo monitoring (1.8%), (HR 0.17, 95% CI 0.02-0.79, p = 0.04). There was no difference in overall CV AEs (grades 3-5) or OS between monitored and unmonitored patients. In the entire cohort, patients with at least one elevated troponin I during the follow up period, during routine monitoring or unmonitored, had a higher risk of overall CV AEs (HR 10.96, 95% CI 4.65–25.85, p<0.001) as well as overall mortality (HR 2.67, 95% CI 1.69 – 4.10, p<0.001) compared to those without elevated troponin. Oncologic time-to-treatment failure (TTF) was not significantly different in a sub-cohort of monitored vs. unmonitored patients.

**Conclusions:** Patients undergoing cardiotoxicity screening with troponin I monitoring during ICI therapy had a lower rate of severe (grade 4-5) CV AEs compared patients who were not screened. Troponin I elevation in screened and unscreened patients was significantly associated with increased CV AEs as well as increased mortality. Troponin I monitoring did not impact oncologic time-to-treatment-failure in a sub-cohort analysis of patients treated with ICI. These results provide preliminary evidence for clinical utility of cardiotoxicity screening with troponin I monitoring in patients receiving ICI therapy.

## Introduction

Immune checkpoint inhibitors (ICI) are a potent and effective form of oncologic therapy, with rapidly expanding use across different malignancies^1^. However, ICIs can cause a wide array of immune-related adverse events (irAEs)^2^, including fulminant myocarditis. Although uncommon, with early studies quoting an incidence between 0.08-1.4%, ICI myocarditis can have a mortality rate estimated up to 40%^2–4^. In addition to myocarditis, ICI therapy has also been associated with increased rates of pericardial disease, cardiac arrythmias, heart failure, ischemic disease, and sudden cardiac death^5–8^.

Given the high rates of cardiac adverse events in patients receiving ICI therapy, there have been several strategies proposed for early detection and screening in these patients^5,9,10^. One such proposal is screening with serial serum cardiac troponins. Elevated levels of both cardiac troponin I and T have been associated with variety of cardiac pathologies, including myocarditis^2,10,11^. Furthermore, elevated serum troponin T at the time of initial disease presentation has been found to be associated with increased mortality in ICI myocarditis^12^. The NCCN guidelines version 2.2023 for Management of Immune Checkpoint Inhibitor-Related Toxicities has a category 2A recommendation to consider high-sensitivity troponin and NT-proBNP at baseline and serially during treatment for those at increased risk. The current European Society of Cardiology guidelines also recommend measurement of baseline cardiac troponin (class I) and consideration of regular troponin monitoring during initial therapy with doses 2, 3, and 4 followed by with every three doses until completion of therapy (class IIa)^5^.

However, there is a paucity of data to guide definitive recommendations regarding the routine use of proactive, serial troponin monitoring in asymptomatic patients on ICI therapy, and practice patterns remain heterogenous across centers^13^. Additionally, opinions remain mixed regarding the universal implementation of such monitoring, with concerns that this could lead to a high rate of false positives, excessive follow-up testing, and possible delays in oncologic therapy^14,15^.

Our group initially piloted a troponin I based screening protocol in patients at Stanford Health Care receiving ICI therapy to detect early cases of ICI cardiotoxicity and allow earlier intervention in cases with potential downstream cardiac morbidity/mortality^10^. We found that routine, serial monitoring of troponin I at baseline and with each dose of ICI in all patients on immunotherapy detected earlier cases of subclinical and asymptomatic ICI myocarditis, prior to onset of more serious cardiac complications such as reduced LVEF, ventricular arrhythmias or sudden cardiac death^10^. The implementation of this screening protocol was made feasible by a rapid cardio-oncology response team triaging elevated troponin values to avoid superfluous testing while catching probable or definite cases by Bonaca criteria^10,16^. Since then, several other studies have found that troponin screening led to higher detected rates of ICI-related cardiac toxicities^17–19^.

Here, we provide an extension of our prior investigations studying the impact of routine troponin I screening in all patients undergoing ICI therapy. The objective of this study was to 1) evaluate the association between cardiotoxicity screening using serial troponin I monitoring and cardiac outcomes and overall survival in patients receiving ICI therapy and 2) assess the predictive value of an elevated on-treatment troponin I for cardiac outcomes. We hypothesized that troponin I monitoring would be associated with improved cardiac outcomes, given that screening may lead to earlier detection of cardiotoxicity and intervention in these patients.

## Methods

We performed an analysis of data collected from 825 patients at Stanford Health Care (SHC) who received ICI therapy between January 2018 and October 2021. Our study included two temporally separated groups: a cohort of 428 patients who underwent routine serial troponin monitoring with each dose of ICI (approx. every 2-4 weeks) during a time when all patients on ICI were screened (Jan 2019 – Oct 2021), and a separate cohort of 397 patients who received ICI therapy at SHC prior to the initiation of routine serial troponin monitoring in all patients at our center (Jan-Dec 2018) (Figure 1).

**Figure 1.**
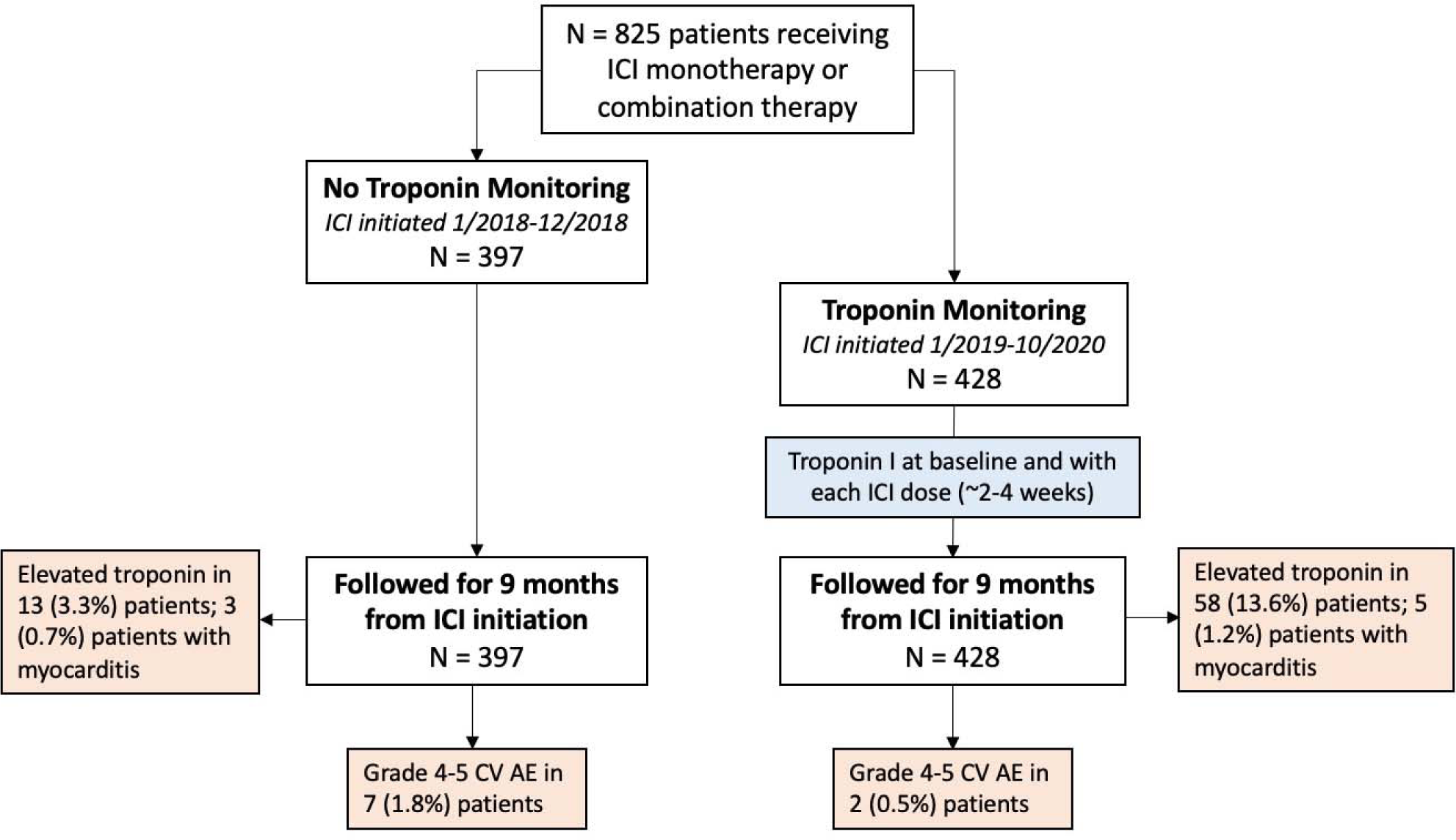
Troponin monitoring algorithm. Abbreviations: ICI = immune checkpoint inhibit

Troponin I monitoring was standardized using an electronic medical record (EMR) lab order set routinely used by oncologists and advanced practitioners to order immunotherapy at our institution. The default lab order template for all patients undergoing ICI therapy included troponin I measurement prior to each ICI dose, including an initial baseline troponin I, unless otherwise opted out by the ordering provider through deselecting the lab on the order set. Opting out could occur as a result of the patient getting labs outside of the Stanford system. As part of this order set, troponin I was measured in blood using Siemens Dimension-EXL (Siemens, Newark, Delaware). Elevated troponin I measurement was defined as ≥55 ng/l and negative measurement as <55 ng/l (the 99th percentile concentration cut-off for the general population defined by the lab assay). Troponin T or high sensitivity-troponin I from other vendors at the time of monitoring were excluded. Patients were included in the troponin monitoring group if they received at least one dose of ICI therapy during a specified period (Jan 2019 – Oct 2021) and had at least one troponin I measured at baseline and after receiving ICI therapy.

The control cohort was identified through the STAnford Research Repository (STARR), an online tool containing medical record data from Stanford Hospital and Clinics. The cohort included a randomly selected group of patients who received at least one dose of ICI therapy during a specified period of January 2018 to December 2018, approximately matched in size with the monitored cohort. This period was chosen to occur immediately before the first dose of ICI was received by patients in the monitored cohort (January 2019) to minimize confounding factors which could occur as a result of the time period shift between the two cohorts. Identical criteria and troponin I thresholds were applied in the control cohort, including an elevated troponin I level defined as troponin I ≥55 ng/l (>99^th^ percentile of the general population). Patients without elevated troponin I measurement included both patients with a negative troponin I measurement and patients with no troponin I measured after ICI initiation and before the end of the follow up period.

Primary outcomes included overall survival (OS) and incidence of cardiovascular adverse events (CV AEs). CV AEs were defined according to the National Cancer Institute (NCI) Common Terminology Criteria for Adverse Events version 5 (CTCAE v5) and included grade ≥ 3 CV AEs of tachy- and bradyarrhythmia, myocarditis, congestive heart failure, acute coronary syndrome, stroke and pericardial effusion. These outcomes were chosen based on their association with ICI as irAEs emerging in the literature.^5,6^ We also identified cases of severe CV AEs, defined as grade 4-5 CV AEs of tachy- and bradyarrhythmia, myocarditis, congestive heart failure, acute coronary syndrome, stroke and pericardial effusion, resulting in critical illness requiring intensive care or death. In patients with more than one CV AE in the follow up period, we only included first CV AE to occur. These events were initially obtained through a search in STARR, with confirmation of each event performed through individual patient chart review in the EMR.

We retrieved events which occurred within nine months of each individual patient’s first dose of ICI for all patients. Nine month follow up was chosen given the latest ICI start date in the monitoring cohort was October 2021, and data collection and analysis was July 2022.

To investigate whether detection of elevated troponin through monitoring might have prompted early discontinuation of ICI therapy, we aimed to compare oncologic time-to-treatment-failure (TTF) between monitored and unmonitored patients. We chose to assess TTF in a subgroup of patients with metastatic melanoma, given that TTF data was not readily available in other subgroups. TTF was defined as the interval from ICI initiation to discontinuation for any reason.

### Statistical Analysis

We used Cox proportional hazards regression to evaluate the association between troponin monitoring and the primary outcomes of overall survival and time to CV AEs. The variables included in multivariable models included age, gender, ICI monotherapy vs dual ICI therapy, combination of chemotherapy and ICI, chemotherapy prior to ICI, and history of diabetes, hypertension, congestive heart failure, coronary artery disease or stroke in the initial 9-month cohort. Given the low incidence of each individual CV AE outcome (tachy- and bradyarrhythmia, myocarditis, congestive heart failure, acute coronary syndrome, stroke and pericardial effusion), we analyzed time to onset of CV AEs collectively and not individually.

We also used Cox regression to analyze the association between the detection of an elevated troponin and the primary outcomes of OS and CV AEs, again adjusting for age, gender, ICI monotherapy vs dual ICI therapy, combination chemotherapy with ICI, chemotherapy prior to ICI, and history of diabetes, hypertension, congestive heart failure, coronary artery disease/stroke. We additionally performed Kaplan-Meyer analysis to compare CV AE-free survival between patients with normal and elevated troponins.

We used Kaplan-Meyer analysis to compare TTF between monitored and unmonitored patients with melanoma.

Analyses were done using GraphPad Prism 9 statistical software. Stanford University Institutional Review Board approved our research protocol. We considered a two-tailed *P*<0.05 to be statistically significant.

## Results

Sociodemographic features, cardiac risk factors and co-morbidities, cancer therapy history, and cancer sub-type were tabulated by exposure to troponin monitoring and the groups were compared using a chi-squared test (Table 1). We included dual ICI therapy, combination chemotherapy with ICI therapy, and prior cardiovascular disease in our analysis given these factors have been associated with higher rates of ICI related CV toxicity ^20,21^.

**Table 1.** Baseline Characteristics of Patient Cohort and Comparison Between Monitored and Unmonitored Groups Values are shown as mean (SD) or n (%). Other types of cancers included: adrenal cancer, basal cell carcinoma, cholangiocarcinoma, gastro-esophageal cancer, germ cell tumor, Hodgkin’s lymphoma, mesothelioma, Merkel cell carcinoma, pancreatic cancer, prostate cancer, small cell lung cancer, squamous cell carcinoma of the skin, thyroid cancer. Abbreviations: CAD = coronary artery disease. ICI = immune checkpoint inhibitors

| **Table 1**<br>Baseline Characteristics of the Study Groups |  |  |  |
| --- | --- | --- | --- |
|  | No Troponin Monitoring<br>(n = 397) | Troponin Monitoring<br>(n = 428) | p Value |
| Male | 227 (57.2%) | 258 (60.2%) | 0.37 |
| Age (years) | 67.06 (12.94) | 67.02 (14.57) | 0.79 |
| CAD | 85 (21.4%) | 94 (22%) | 0.85 |
| Stroke | 18 (4.5%) | 32 (7.5%) | 0.08 |
| Diabetes Mellitus | 67 (16.9%) | 85 (19.8%) | 0.27 |
| Hypertension | 224 (56.4%) | 227 (53%) | 0.33 |
| Congestive Heart Failure | 35 (8.8%) | 26 (6.1%) | 0.13 |
| Arrhythmia | 67 (16.8%) | 59 (13.8%) | 0.22 |
| Cytotoxic Chemotherapy Received |  |  |  |
| Chemo Prior to ICI | 209 (52.6%) | 220 (51.4%) | 0.72 |
| Chemo Concurrent with ICI | 61 (15.4%) | 101 (23.6%) | 0.01 |
| Malignancy Type |  |  |  |
| Non-small Cell Lung Cancer | 115 (29%) | 103 (24.1%) | 0.11 |
| Melanoma | 45 (11.3%) | 44 (10.2%) | 0.62 |
| Head and Neck Cancer | 42 (10.6%) | 44 (10.3%) | 0.89 |
| Renal Cell Carcinoma | 32 (8.1%) | 52 (12.1%) | 0.05 |
| Breast Cancer | 14 (3.5%) | 12 (2.8%) | 0.55 |
| Urothelial and Bladder Cancer | 25 (6.3%) | 45 (10.5%) | 0.04 |
| Gynecologic Cancer | 19 (4.8%) | 24 (5.6%) | 0.60 |
| Sarcoma | 19 (4.8%) | 18 (4.2%) | 0.69 |
| Hepatocellular Carcinoma | 14 (3.5%) | 9 (2.1%) | 0.21 |
| Colorectal and Anal Cancer | 11 (2.8%) | 10 (2.3%) | 0.69 |
| Other | 61 (15.4%) | 66 (15.4%) | 0.98 |
| ICI Type |  |  |  |
| Anti PD-1 | 297 (74.8%) | 318 (74.3%) | 0.87 |
| Anti PD-L1 | 46 (11.6%) | 54 (12.6%) | 0.65 |
| Anti CTLA-4 | 3 (0.7%) | 0 (0%) | 0.07 |
| Anti CTLA-4 + PD-1 | 51 (12.8%) | 56 (13.1%) | 0.92 |

Of the 825 participants, 428 (52%) were evaluated through the standard troponin I monitoring protocol, and 397 (48%) patients belonged to the unmonitored group prior to onset of routine monitoring (Figure 1). The mean (SD) age of the full cohort was 66.9 (13.8), and 485 (58.6%) were male. There were no significant differences between the two groups with regards to gender, age, baseline comorbidities and ICI type (Table 1). There was a significantly higher proportion of patients receiving chemotherapy in combination with ICI therapy in the troponin monitoring group (23.6% vs 15.4%, p = 0.003). Additionally, the patients who underwent troponin monitoring had a significantly higher proportion of patients being treated for urothelial and bladder cancer (10.5% vs 6.3%, p = 0.039).

There were 82 patients (20%) in the unmonitored group with any troponin I measured during the nine month follow up period; most elevated troponin I measurements in the unmonitored group occurred in the setting of emergency department visits or hospital admission.

There were 71 patients (8.6%) with an elevated troponin I measurement during the nine-month follow-up period. The total incidence of elevated troponin was 58/428 (13.6%) in the monitored group and 13/397 (3.3%) in the unmonitored group (Figure 1). In all patients, those with an elevated troponin I had a higher risk of CV AEs compared to those without elevated troponin measurements (HR 10.96, 95% CI 4.65–25.85, p<0.001). This finding remained consistent when evaluated in the monitored group alone (HR 8.64, 95% CI 3.04 – 25.38, p<0.001) (Figure 4). Additionally, patients with an elevated troponin I during the study period had higher risk of all-cause mortality, seen in all patients (HR 2.67, 95% CI 1.69 – 4.10, p<0.001) as well as monitored patients alone (HR 1.93, 95% CI 1.09 – 3.25, p = 0.02) (Table 3, Figure 4).

**Figure 4.**
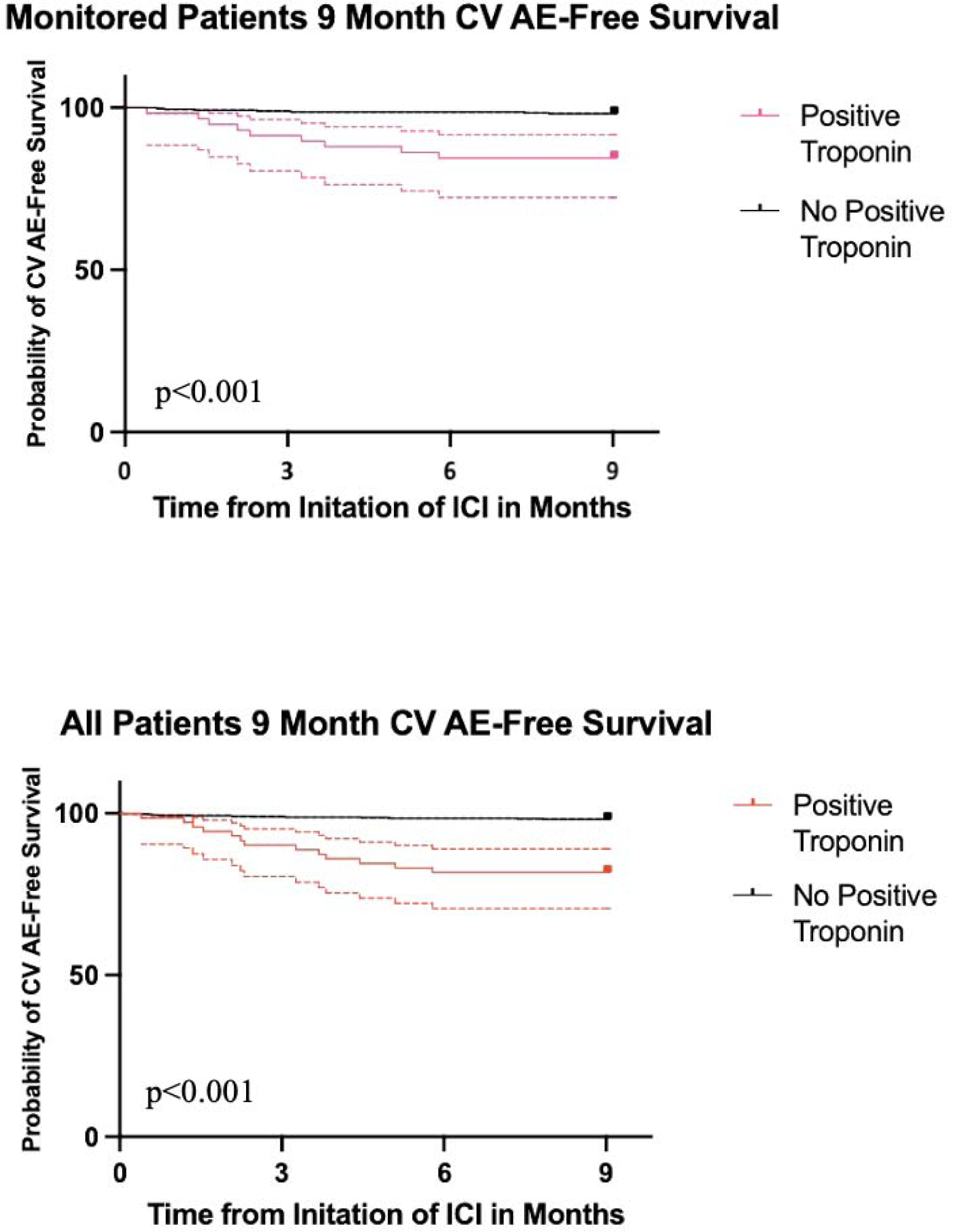
Kaplan Meier analysis comparing CV AE-Free survival between patients with elevated and normal troponin, with 95% confidence interval. Abbreviations: CV AE = cardiovascular adverse events, OS = overall survival.

**Table 3.** Association between Elevated Troponin and Outcomes of Death and Grade ≥ 3 CV AEs in Full Cohort and Monitored Group Values are shown as n (%). Abbreviations: CV AE = cardiovascular adverse event, OS = overall survival

| **Table 3**<br>Association between Elevated Troponin and Outcomes of Death and Grade $\geq 3$ CV AEs in Full Cohort and Monitored Group | | | | | | |
| --- | --- | --- | --- | --- | --- | --- |
| | Grade $\geq 3$<br>CV AEs | Multivariable HR<br>(95% CI) | *P* value | Death | Multivariable HR<br>(95% CI) | *P* value |
| Full Cohort<br>(N = 825) | 27 (3.3%) |  |  | 151 (18.3%) |  |  |
| Normal Troponin<br>(N=755) | 16 (2.1%) | 1 (Reference) |  | 123 (16.3%) | 1 (Reference) |  |
| Elevated Troponin<br>(N=71) | 11 (15.5%) | 10.95 (4.65-25.85) | <0.0001 | 28 (39.4%) | 2.67 (1.69-4.10) | <0.0001 |
| Monitored Cohort<br>(N = 428) | 16 (3.7%) |  |  | 80 (18.7%) |  |  |
| Normal Troponin<br>(N=370) | 7 (1.9%) | 1 (Reference) |  | 61 (16.5%) | 1 (Reference) |  |
| Elevated Troponin<br>(N=58) | 9 (15.5%) | 8.64 (3.04-25.38) | <0.0001 | 19 (32.8%) | 1.93 (1.09-3.25) | 0.02 |

Two patients (0.5%) in the monitored group and seven patients (1.8%) in the unmonitored group experienced severe CV AEs (grade 4-5) (Table 2). There was a significantly lower rate of severe (grade 4-5, requiring intensive care) CV AEs in patients who underwent troponin monitoring compared to patients who were not monitored (HR 0.17, 95% CI 0.02-0.79, p = 0.04) (Table 2). There were two patients who experienced grade 5 CV AE (resulting in death), both in the unmonitored group, including one with severe myocarditis who developed severely reduced left ventricular dysfunction. The incidence of myocarditis was 5/428 (1.2%) in the monitored group, with no severe (grade 4-5) cases, and 3/397 (0.7%) in the unmonitored group, with all three of these cases (100%) being severe (grade 4-5) (Table 2).

**Table 2.** Association between Troponin Monitoring and Outcomes of Death and Severe CV AEs Values are shown as n (%). Hazard ratios represent outcomes in monitored group relative to unmonitored group. Abbreviations: CV AE = cardiovascular adverse event, OS = overall survival

| **Table 2**<br>CV AEs and OS within 9 Months of ICI initiation |  |  |  |  |
| --- | --- | --- | --- | --- |
|  | No Troponin Monitoring<br>(n = 397) | Troponin Monitoring<br>(n = 428) | Multivariable Hazard Ratio<br>(95% CI) | P value |
| Death | 71 (17.9%) | 80 (18.7%) | 1.03 (0.75 - 1.43) | 0.84 |
| Severe CV AEs (grade 4-5) | 7 (1.8%) | 2 (0.5%) | 0.17 (0.02 – 0.79) | 0.04 |
| Arrhythmia | 3 (0.7%) | 2 (0.5%) | - | - |
| Myocarditis | 3 (0.7%) | 0 (0%) | - | - |
| Congestive Heart Failure | 0 (0%) | 0 (0%) | - | - |
| Acute Coronary Syndrome | 0 (0%) | 0 (0%) | - | - |
| Stroke | 1 (0.2%) | 0 (0%) | - | - |
| Pericardial Effusion | 0 (0%) | 0 (0%) | - | - |
| All CV AEs (grade 3-5) | 11 (2.8%) | 16 (3.7%) | 1.34 (0.62 - 3.00) | 0.47 |
| Arrhythmia | 4 (1%) | 2 (0.4%) | - | - |
| Myocarditis | 3 (0.7%) | 5 (1.2%) | - | - |
| Congestive Heart Failure | 1 (0.2%) | 4 (0.9%) | - | - |
| Acute Coronary Syndrome | 1 (0.2%) | 2 (0.4%) | - | - |
| Stroke | 1 (0.2%) | 1 (0.2%) | - | - |
| Pericardial Effusion | 1 (0.2%) | 2 (0.4%) | - | - |

From the time of ICI initiation, the nine-month mortality rate was 18.7% (n=80) in the monitored group and 17.9% (n=71) in the unmonitored group, with no difference in nine-month overall survival (OS) between the two groups (HR 1.03, 95% CI 0.75-1.43, p=0.84) (Figure 2). Incidence of diagnosed grade 3-5 CV AEs was 3.7% (n=16) in the monitored group and 2.8% (n=11) in the unmonitored group, with no difference in incidence of overall diagnosed CV AEs between the two groups (HR 1.34, 95% CI 0.62-3.00, p=0.47) (Table 2).

**Figure 2.**
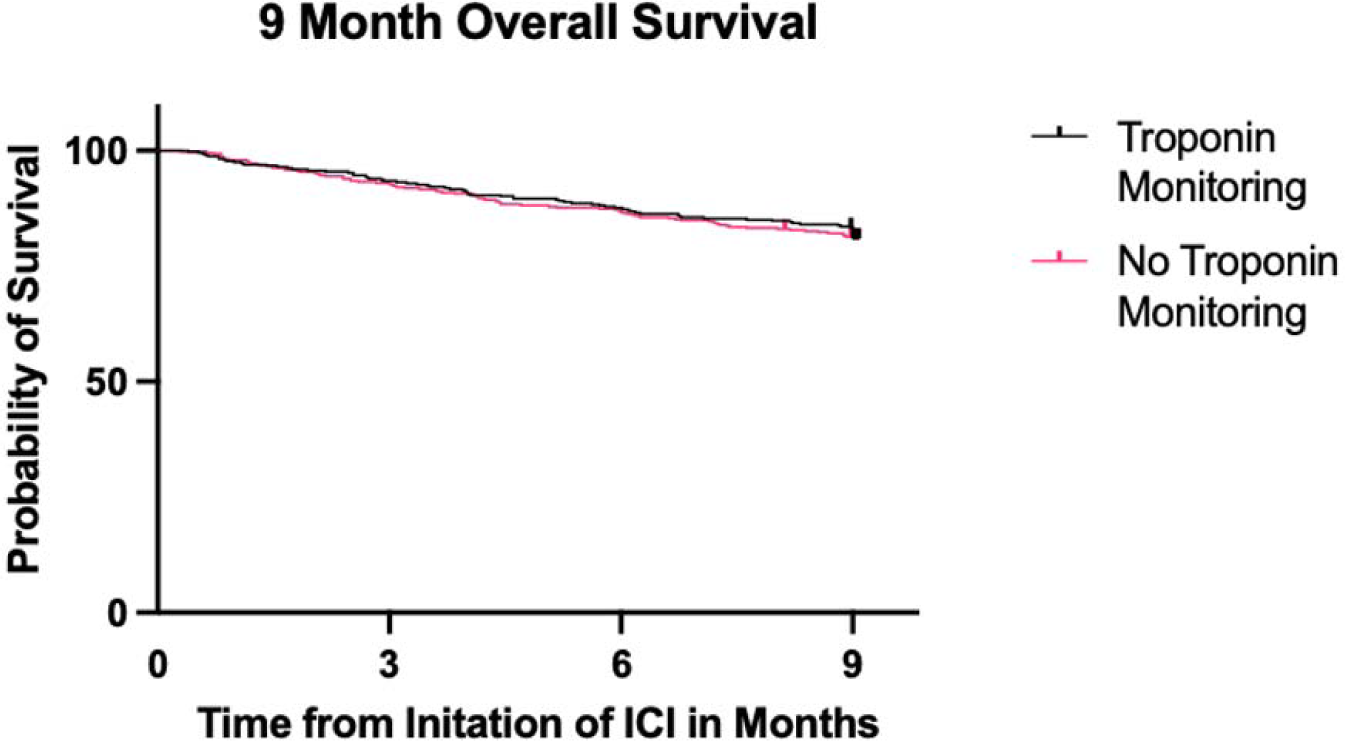
Kaplan-Meier analyses of association between troponin monitoring and overall survival. Abbreviations: ICI = immune checkpoint inhibitor

To investigate oncologic time-to-treatment-failure (TTF), we analyzed a subgroup of patients with metastatic melanoma (n=89), where we found no difference in TTF at nine months between the two groups (p = 0.91), with 24 (54%) monitored patients and 24 (53%) unmonitored patients having discontinued ICI by nine months (Figure 3).

**Figure 3.**
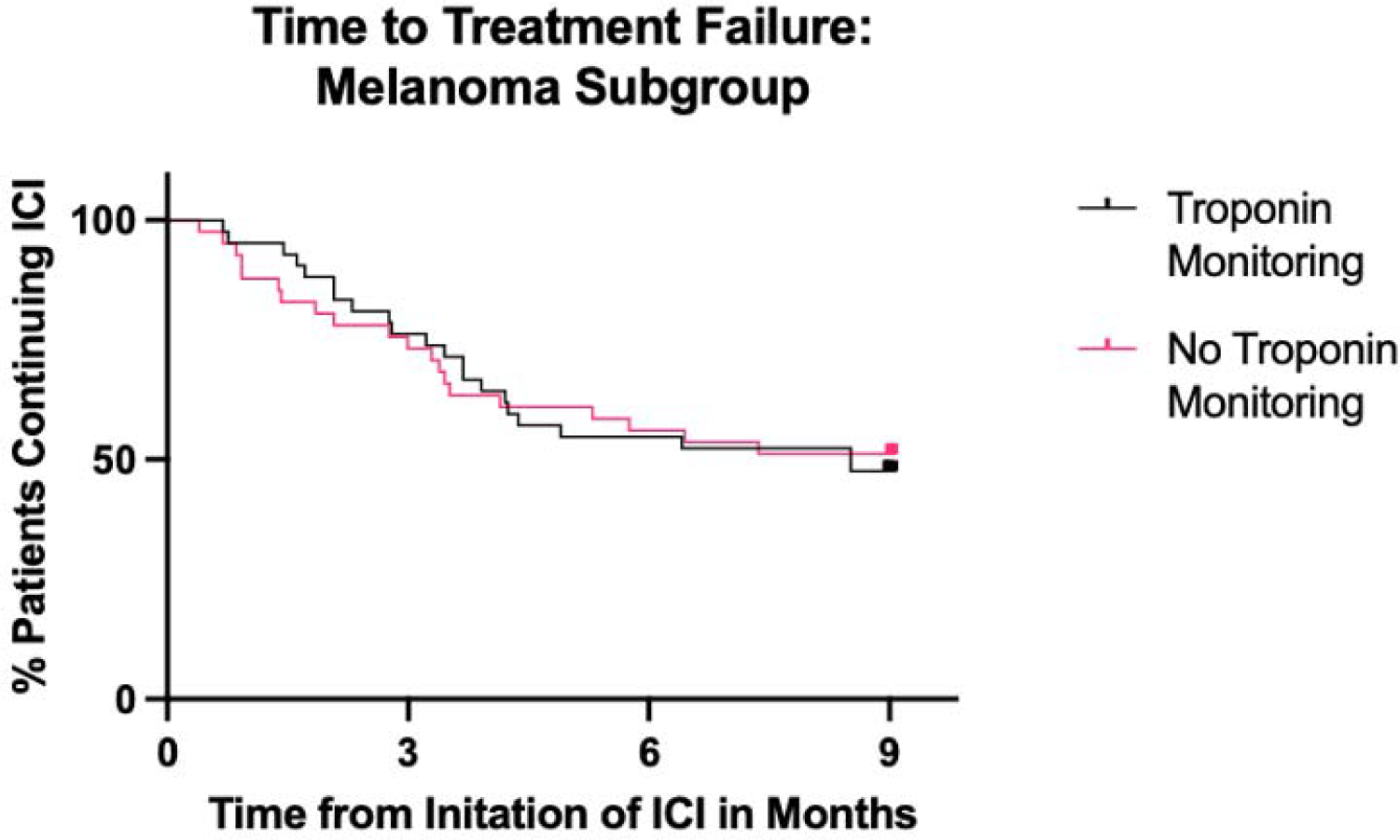
Comparison of Time to Treatment Failure between Troponin Monitored and Unmonitored Groups. Abbreviations: ICI = immune checkpoint inhibitor

## Discussion

This is the largest study to date to evaluate outcomes associated with cardiotoxicity screening with standardized, routine troponin I measurements in all patients receiving immunotherapy at a single center. Our data was prospectively collected, and we compared 428 troponin I monitored patients on ICI during a defined study period to 397 unmonitored patients who underwent ICI therapy in a similarly defined time period immediately prior to routine initiation of monitoring at our center. We found that patients who underwent routine troponin I monitoring at baseline and every dose of ICI had a lower rate of severe CV AE (grade 4-5, resulting in critical illness or death) compared to unmonitored patients. One explanation for this finding is that monitoring troponin I levels may lead to earlier detection and intervention of cardiac toxicity prior to development of more severe toxicity. This hypothesis is supported by the greater number of grade 3 CV AEs in the monitored group compared with the unmonitored, indicating that while there were similar rates of CV AEs between the two groups, the monitored group appeared to have fewer severe events and downstream cardiovascular sequelae. It is important to note that at our center, patients with elevated troponin I levels detected through the monitoring protocol were evaluated by our rapid cardio-oncology triage team, which guided early management of these elevated troponin I levels and aided with proper interpretation of these values in their clinical context and management^10^.

Our detected rate of myocarditis was 1.2% in the monitored group and 0.7% in the unmonitored group, which is consistent with prior literature^2–4^. All patients in the monitored group had grade 3 myocarditis, while the unmonitored group had two patients with grade 4 necessitating intensive care and one patient with grade 5 myocarditis leading to death, in line with our findings of more severe CV AE in the unmonitored patients. This suggests that, although troponin I monitoring did not appear to impact overall survival in the 9-month study period, it did impact degree of cardiovascular morbidity and quality-of-life outcomes in patients who had immune-mediated cardiac events. The fact that we did not observe a difference in overall survival between the two groups in the 9-month long study period could be attributable to the fact that the study period was relatively short and the overall event rate during this period was low. Further studies are needed to study the long-term impacts of troponin monitoring and their impact on patient outcomes.

Our findings support troponin I as a predictive biomarker for mortality in this patient population. Consistent with our findings, prior studies have also demonstrated increased rates of CV AEs in patients undergoing ICI therapy with elevated troponin levels^10,18,19^. However, prior studies have generally measured troponin levels at the time of myocarditis diagnosis, and to our knowledge, no prior study has compared routine use of prospective troponin I screening in all patients undergoing ICI therapy to unmonitored patients. Additionally, we found that prospective screening with elevated troponin I was also associated with increased all-cause mortality. This supports the growing body of evidence that elevated troponin I and T may become useful biomarkers capable of predicting significant adverse outcomes in patients receiving ICI therapy^22^.

Some concerns have been raised that routine troponin monitoring could lead to premature cessation of oncologic therapy and impact cancer survival^14,15^. While cancer outcomes were not a primary outcome of this study, we found no difference in OS between monitored and unmonitored patients at both nine months and two years. Additionally, we were able to obtain oncologic time to treatment failure (TTF) in patients with metastatic melanoma and found no difference between monitored and unmonitored patients, suggesting monitoring did not lead to premature cessation of ICI therapy. Further studies are needed to better understand the impact of troponin monitoring on cancer-related outcomes.

Our study has several limitations. It is a single-center study at a large tertiary care center, which limits the generalizability of our results to other institutions. Our study only included troponin I, which at the time of the study was the only widely used troponin-based assay at our institution, and may not be automatically generalizable to other troponin assays such as troponin T. The availability of immediate cardiac-oncology consultation for elevated troponin was important for interpreting elevated troponin levels and next steps and may not be as readily available in other practice settings. Additionally, the retrospective design of our study and lack of prospective randomization between monitored and unmonitored groups introduces a possible source of bias, as practice variations among individual oncologists may result in patients with higher risk of cardiac events getting more troponin I monitoring. This risk was minimized by incorporating the troponin I assay into a standardized immunotherapy order set and making it an “opt-out” rather than an “opt-in” option used by all oncologists and advanced practitioners at our institution. In an additional attempt to mitigate bias for monitoring of higher risk patients, we chose to use a control group of patients who received ICI therapy prior to the incorporation of troponin into the EMR order set, during a period when no troponins were routinely drawn for monitoring in asymptomatic patients. However, this does create a temporal difference between our two cohorts, during which ICI therapy indications, FDA approvals, and protocols may have changed. These risks related to temporal differences were minimized by choosing time periods immediately adjacent to each other.

Most comorbidities and cardiovascular risk factors were similar between the monitored and unmonitored groups, but there were some differences. There was a higher rate of patients with urothelial cancer in the monitored group, which may be explained in part due to changes in standard of care therapy for patients with solid tumors over time, given the temporal separation of our two cohorts. Additionally, our monitored group had a greater proportion of patients receiving concurrent chemotherapy with ICI, potentially due to a concern from the ordering oncologists for higher risk of cardiac toxicity from concurrent chemotherapy/ICI. We attempted to address relevant baseline differences by doing a multivariable analysis, accounting for relevant comorbidities and type of anti-cancer therapy.

To our knowledge, this is the largest cohort analysis investigating outcomes associated with routine serial troponin monitoring in all patients undergoing ICI therapy at a single center and the first to compare outcomes between monitored and unmonitored patients. Our population was heterogenous in terms of cancer type and types of anti-cancer therapies received. We detected a lower rate of severe (grade 4-5) CV AEs in patients who underwent serial troponin monitoring. Additionally, we found no difference in OS or oncologic TTF between monitored and unmonitored patients. Furthermore, we corroborated recent findings demonstrating the utility of troponin I has a prognostic biomarker, this time in a prospective asymptomatic cohort of patients receiving ICI, rather than at the time of ICI myocarditis diagnosis as was previously demonstrated^12^. These findings suggest that biomarker-based cardiotoxicity screening, when used judiciously in the presence of a cardio-oncology triage team, may be helpful as a prognostic tool as well as in reducing cardiovascular morbidity and preventing more fulminant cardiac toxicities through earlier detection at a lower grade. These findings are suggestive and hypothesis-generating, and further prospective randomized studies are needed to evaluate the long-term impacts of routine biomarker-based screening on cardiovascular and cancer-related outcomes.

## Data Availability

All data produced in the present study are available upon reasonable request to the authors.

## Acknowledgments

We would like to thank the entire Stanford Cardio-Oncology team, including oncologists and cardiologists, who have contributed to the excellent clinical care of the patients in this study and have provided the necessary data for the analysis in the study.

