## Supplementary figures and images for "THE IMPACT OF ROUTINE CARDIAC TROPONIN I-BASED CARDIOTOXICITY SCREENING ON CLINICAL OUTCOMES IN PATIENTS ON CANCER IMMUNOTHERAPY"

### Supplemental Tables

Supplementary Tables:
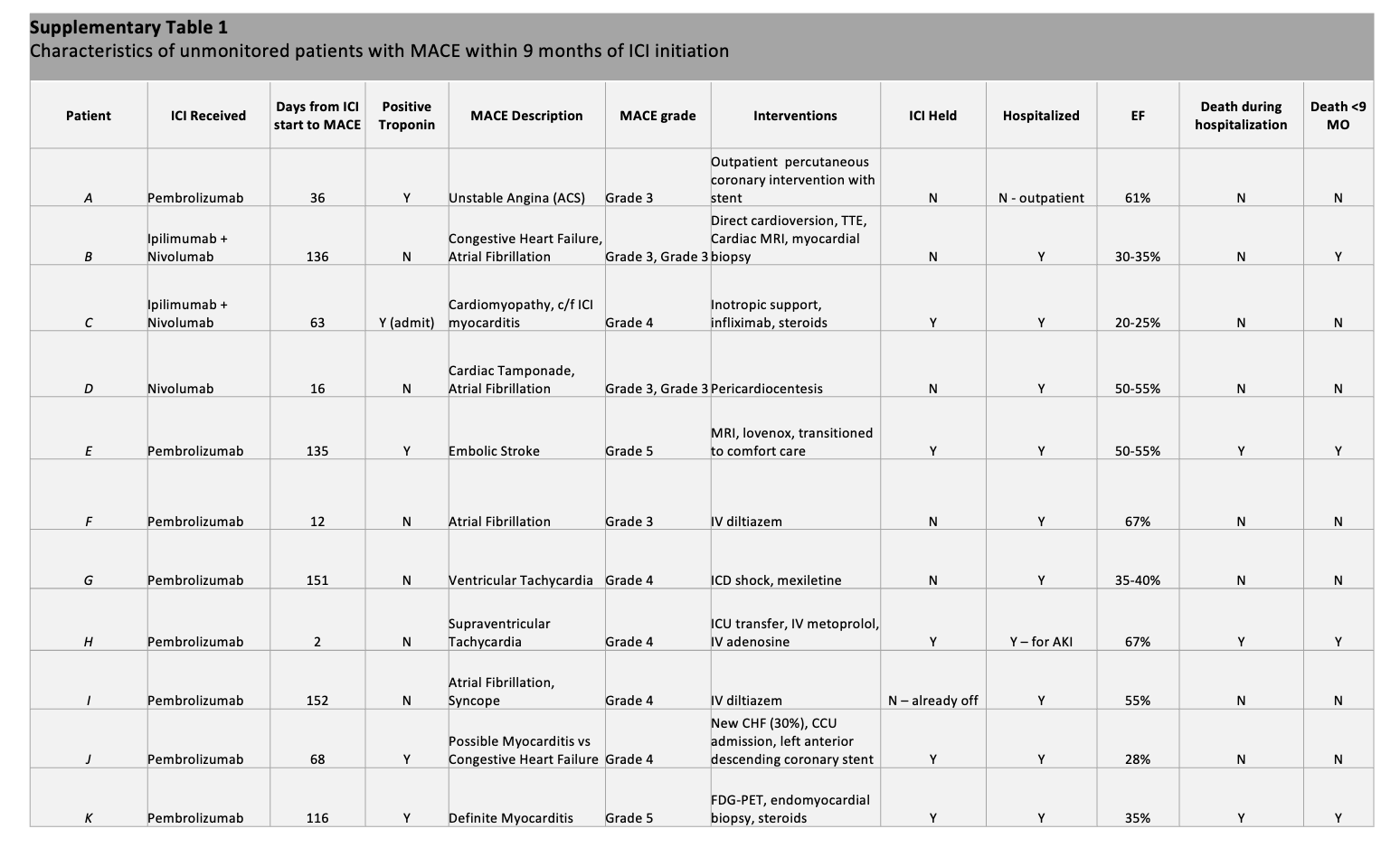

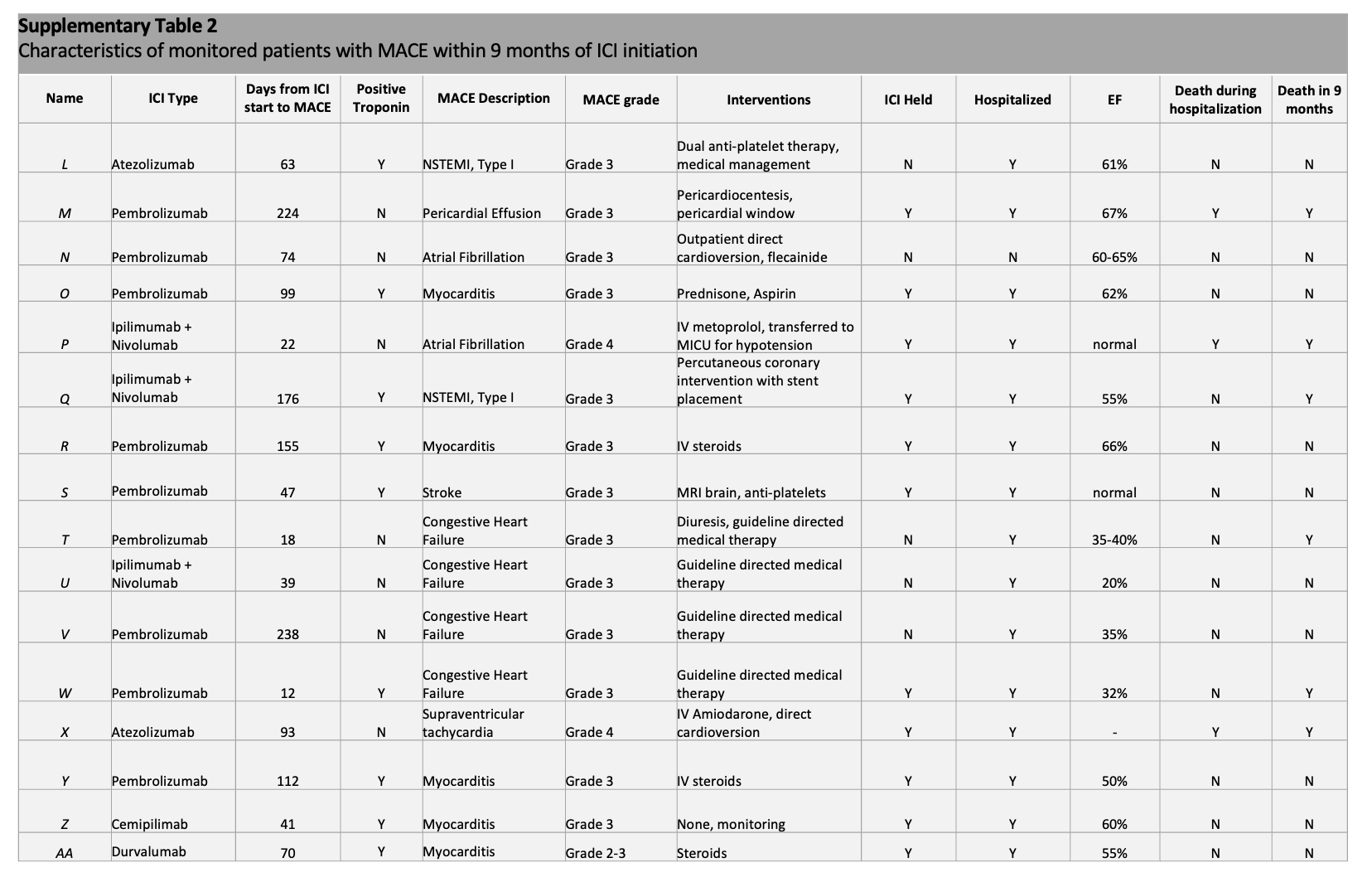
